# Predicting and Explaining WASH Inequities in Nigeria: A Machine Learning Approach to Accelerate NTD Elimination

**DOI:** 10.64898/2026.09.14.26363014

**Authors:** Nneoma Lydia Dorbor, Shitta Kefas Babale, Gbenga Alege

## Abstract

**Background:** Inadequate water, sanitation, and hygiene (WASH) remain key risk factors for neglected tropical diseases (NTDs) in Nigeria. However, the complex, non-linear determinants of household WASH access are poorly understood, limiting targeted NTD control.

**Methods:** We conducted a cross-sectional secondary analysis of 30,045 households from the nationally representative 2024 Nigeria Demographic and Health Survey (NDHS). Four supervised machine learning models — Logistic Regression, Decision Tree, Random Forest, and XGBoost — were trained to predict improved household WASH access. Model performance was evaluated using accuracy, precision, recall, F1-score, and AUC. Explainability was achieved using SHAP values and feature importance.

**Results:** XGBoost achieved the highest performance: 83.3% accuracy, 86.9% precision, 92.3% recall, 89.5% F1-score, and AUC 0.877. Random Forest performed similarly with AUC 0.868. SHAP analysis identified household wealth index, improved sanitation, place of residence, and water availability at handwashing stations as the top predictors. Rural households in northern Nigeria and those in the lowest wealth quintiles had the lowest predicted probability of improved WASH.

**Conclusions:** Explainable machine learning effectively identifies populations most vulnerable to WASH inequities. Targeting low-wealth, rural households in northern Nigeria with integrated WASH interventions could yield the greatest gains for NTD control. Our open-source code provides a reproducible framework for evidence-based resource allocation.

**Author Summary:** Neglected tropical diseases thrive where water and sanitation are poor. In Nigeria, millions still lack access to basic WASH, but programs don’t know exactly which households to target first.

Using data from 30,045 households in the 2024 Nigeria Demographic and Health Survey, we used artificial intelligence to predict who is most likely to lack improved WASH. The XGBoost model was 83% accurate and showed that household wealth, type of toilet, rural location, and water at handwashing stations are the biggest factors.

Our results suggest that NTD programs should focus integrated WASH support on low-wealth, rural households, especially in northern Nigeria. By using this open-source AI tool, Nigeria can move from blanket interventions to precision targeting and accelerate progress toward NTD elimination.

## 1. Introduction

Access to safe water, adequate sanitation, and appropriate hygiene is fundamental to human health and to achieving Sustainable Development Goals 3 and 6. WASH interventions interrupt faecal-oral transmission pathways and are critical for preventing diarrheal diseases and several NTDs, including soil-transmitted helminths and trachoma.[1][17]

Despite progress, WASH access in Nigeria remains deeply unequal across socioeconomic and urban-rural lines. This inequality directly undermines NTD control efforts, as transmission persists in communities with poor WASH infrastructure.[3][8]

Conventional regression methods often miss complex interactions between household factors. Machine learning (ML) offers a complementary approach for classification and prediction where relationships are non-linear. When paired with Explainable AI like SHAP, ML becomes transparent and actionable for public health.[10][11]

This study had 2 aims: 1) Compare Logistic Regression, Decision Tree, Random Forest, and XGBoost for predicting household WASH access using 2024 NDHS data, and 2) Use SHAP to identify the key drivers of WASH inequity to inform NTD control policy in Nigeria.

## 2. Materials and Methods

### 2.1 Study Design and Data Source

Cross-sectional secondary analysis of the 2024 Nigeria Demographic and Health Survey (NDHS) household recode. The NDHS is a nationally representative survey implemented by the National Population Commission and ICF. Data access was approved through the DHS Program.

### 2.2 Study Population and Sample

The analytical sample included 30,045 households with complete data for the outcome and predictors after cleaning.

### 2.3 Outcome Variable

Binary outcome: ‘1 = Improved WASH access‘, ‘0 = Unimproved WASH access‘. Constructed using NDHS standard definitions for improved water, sanitation, and hygiene.

A secondary outcome of predicted probability of unimproved WASH was aggregated to geopolitical zone level for geospatial visualization.

### 2.4 Predictor Variables

Selected based on literature and availability:

- Household wealth index ‘hv270’

- Place of residence ‘hv025’: Urban/Rural

- Improved sanitation facility: derived from ‘hv219’

- Water available at handwashing station: derived from ‘hv230b’

### 2.5 Data Preprocessing

Conducted in Python 3.10 using pandas, scikit-learn. Steps: import, variable selection, duplicate removal, missing value handling, feature scaling, and 80:20 train-test split with random seed 42.

### 2.6 Machine Learning Models

Four classifiers were evaluated: Logistic Regression, Decision Tree, Random Forest, and XGBoost. XGBoost uses gradient boosting with regularization to handle complex interactions.[5]

**Fig. 1:**
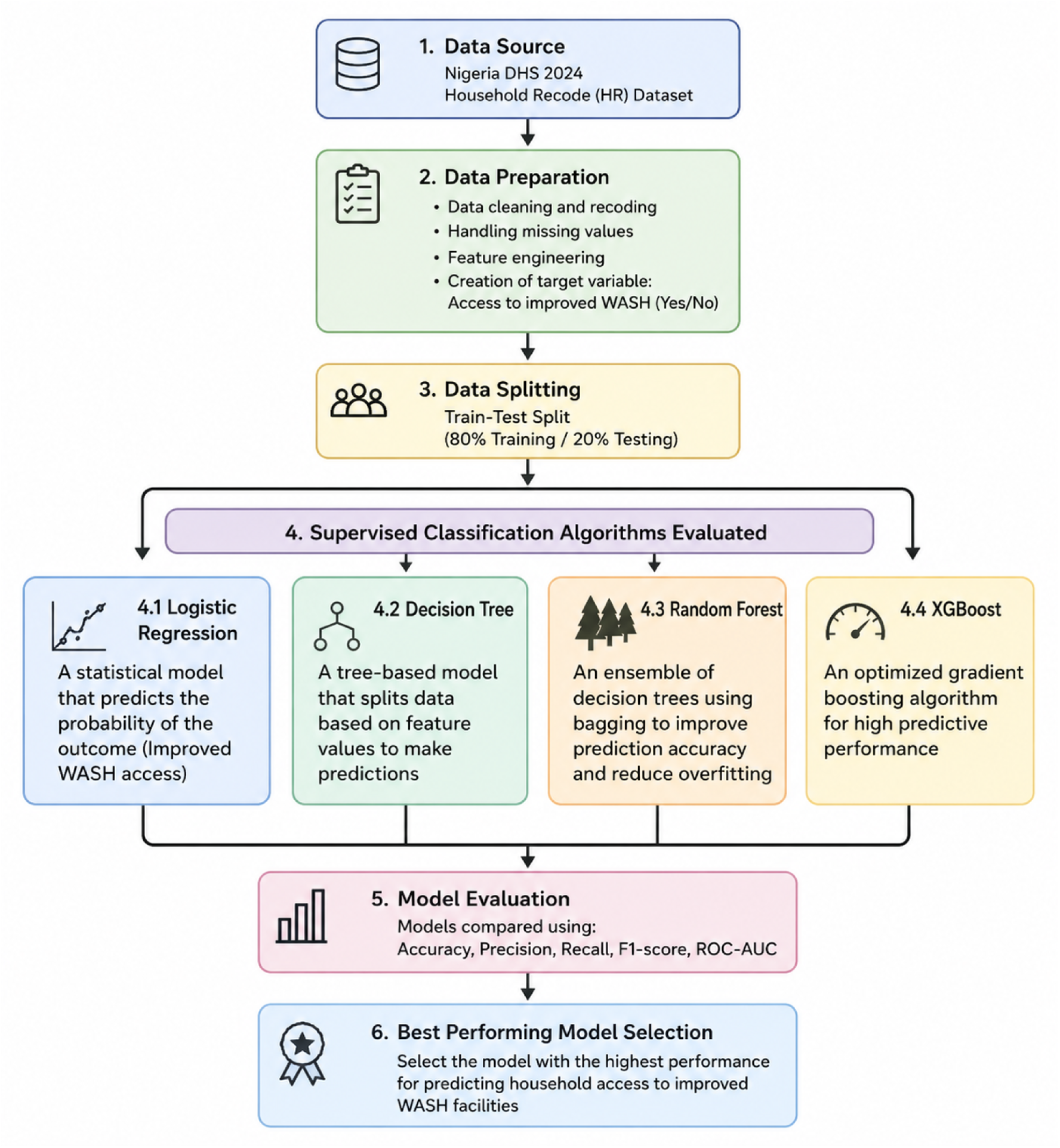
A flowchat explaining the algorithms evaluated

### 2.7 Model Evaluation

Due to class imbalance (77% improved WASH), we used accuracy, precision, recall, F1-score, and AUC. Higher recall is critical for identifying vulnerable households.

### 2.8 Model Explainability

Feature importance and SHAP were applied to the best model to quantify direction and magnitude of each predictor’s contribution.[10][11]

### 2.9 Software and Reproducibility

Python with scikit-learn, XGBoost, SHAP, and Matplotlib. Full code and environment are available at: ‘https://github.com/nneoma-lydia/WASH-NDHS-Nigeria-2024’

### 2.10 Ethical Approval

Used de-identified public DHS data. Original NDHS ethical approval obtained by NPC and ICF.

### 2.11 Geospatial Data

Geopolitical zone boundaries for Nigeria were obtained from the Humanitarian Data Exchange (HDX) GADM Level 1 administrative boundary shapefile for Nigeria [18]. The predicted household-level probability of unimproved WASH was aggregated to the zone level and joined to the shapefile in Python using GeoPandas for choropleth visualization.

## 3. Results

### 3.1 Household Characteristics

N=30,045. 51.5% urban, 48.5% rural. Wealth: 15.6% poorest, 27.1% richest. Urban households had higher rates of improved water (77.5% vs 22.5%), sanitation (66.6% vs 33.4%), and handwashing (69.3% vs 30.7%).

### 3.2 Bivariate Associations

Significant associations with WASH access: Residence χ²=4,121.74, p<0.001; Wealth χ²=8,255.80, p<0.001.

### 3.3 Logistic Regression

Significant predictors: Residence AOR=2.04, Wealth AOR=2.16, Improved toilet AOR=1.96, Handwashing water AOR=1.42. All p<0.001.

### 3.4 Model Performance Comparison

**Table 1.** Performance metrics of machine learning models for predicting improved household WASH access, NDHS 2024. Comparison of 4 supervised ML models on the 20% test set (n=6,009 households). Metrics include accuracy, precision, recall, F1-score, and AUC. XGBoost achieved the highest overall performance with 83.3% accuracy and AUC of 0.877. All models had high recall >89%, indicating strong ability to identify households with improved WASH. Random Forest had the highest precision at 87.0%.

| Model | Accuracy | Precision | Recall | F1-score | AUC |
| --- | --- | --- | --- | --- | --- |
| XGBoost | 83.3% | 86.9% | 92.3% | 89.5% | 0.877 |
| Random Forest | 83.2% | 87.0% | 92.0% | 89.4% | 0.868 |
| Logistic Regression | 80.2% | 84.1% | 90.4% | 87.1% | 0.860 |
| Decision Tree | 79.1% | 82.8% | 89.2% | 85.9% | 0.842 |

Note: AUC = Area Under the Receiver Operating Characteristic Curve.

**Fig 2.**
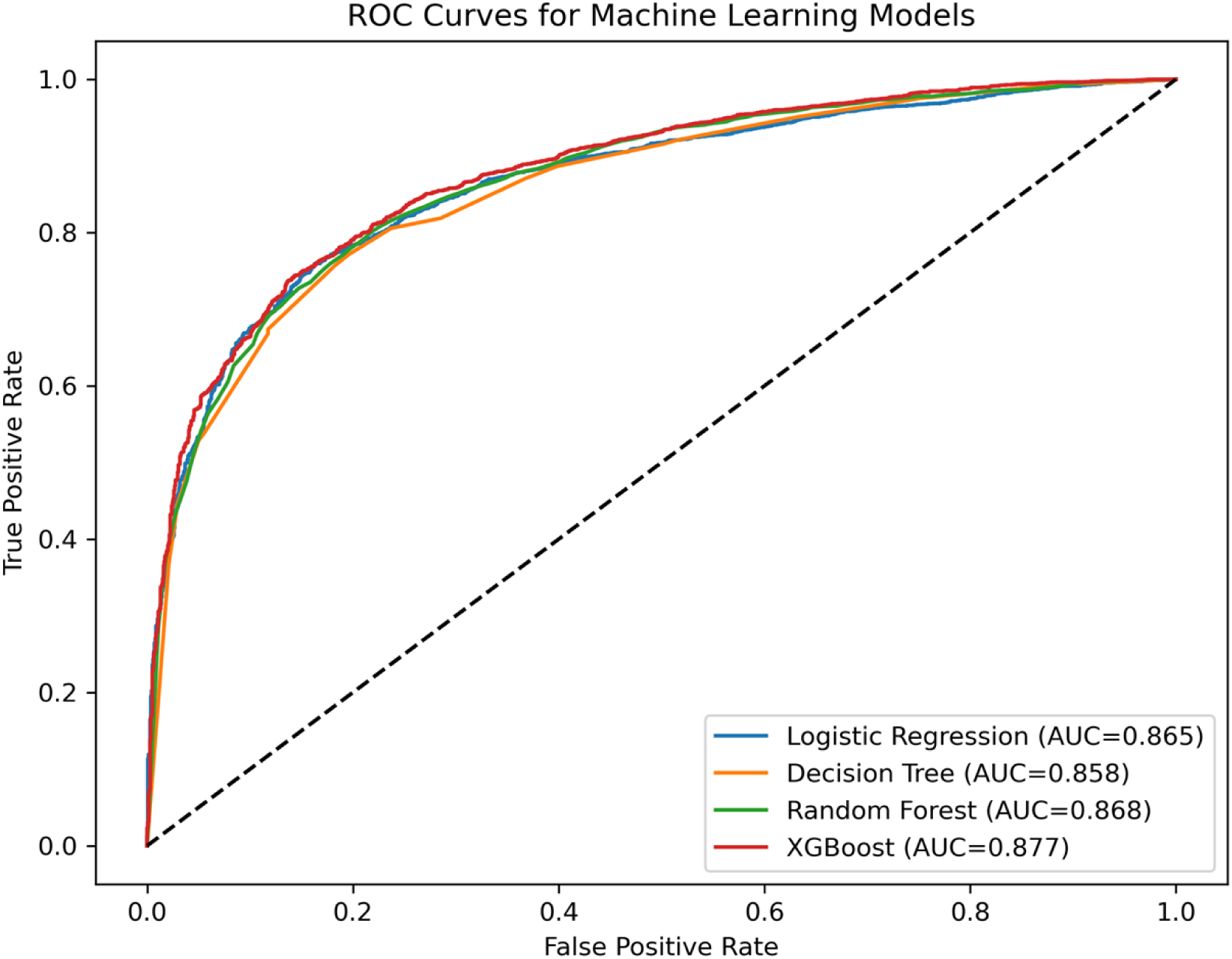
ROC curves for machine learning models predicting improved WASH access. Receiver Operating Characteristic curves comparing Logistic Regression, Decision Tree, Random Forest, and XGBoost. XGBoost achieved the highest AUC of 0.877.

**Table 2.**
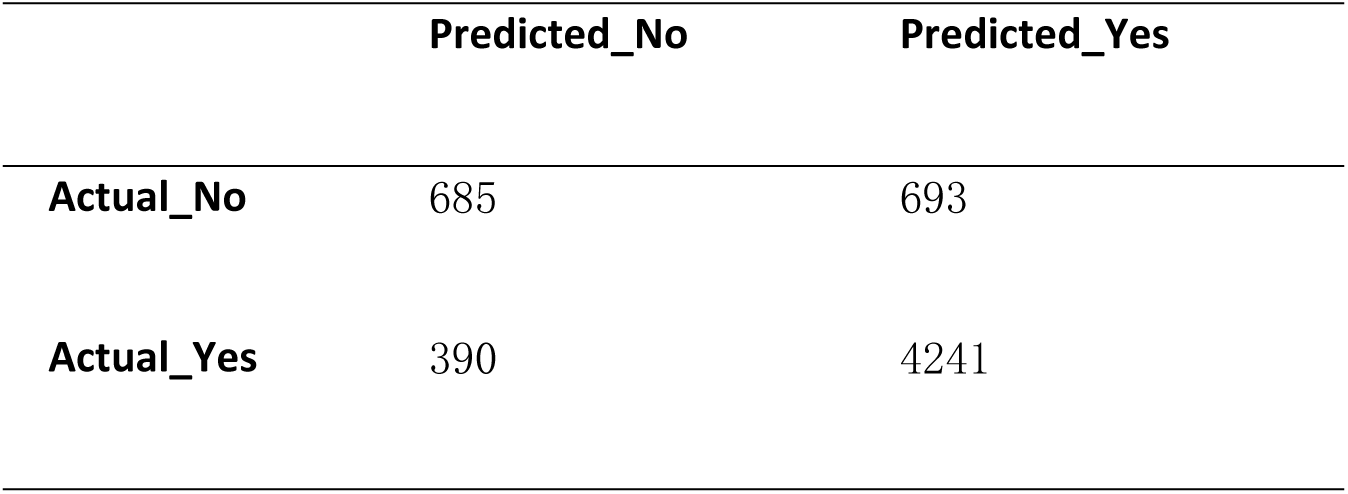
Confusion matrix for Decision Tree model. True vs predicted classes for improved vs unimproved WASH access

**Table 3.**
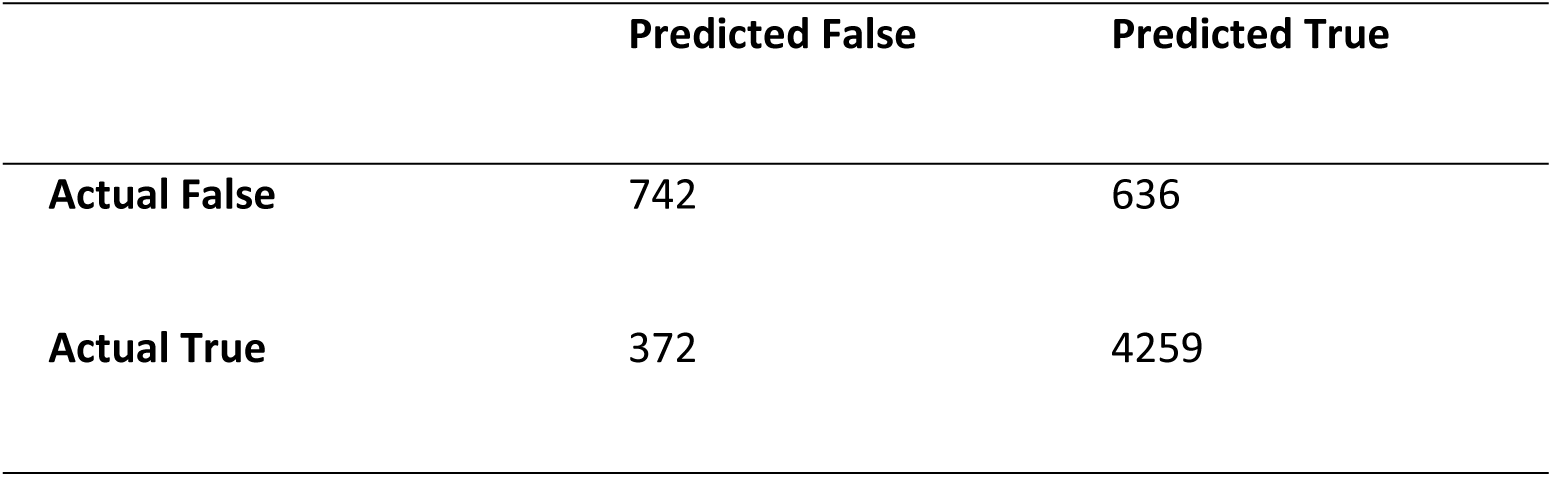
Confusion matrix for Random Forest model. True vs predicted classes for improved vs unimproved WASH access using Random Forest on test data.

|  | Predicted False | Predicted True |
| --- | --- | --- |
| Actual False | 742 | 636 |
| Actual True | 372 | 4259 |

**Fig 3.**
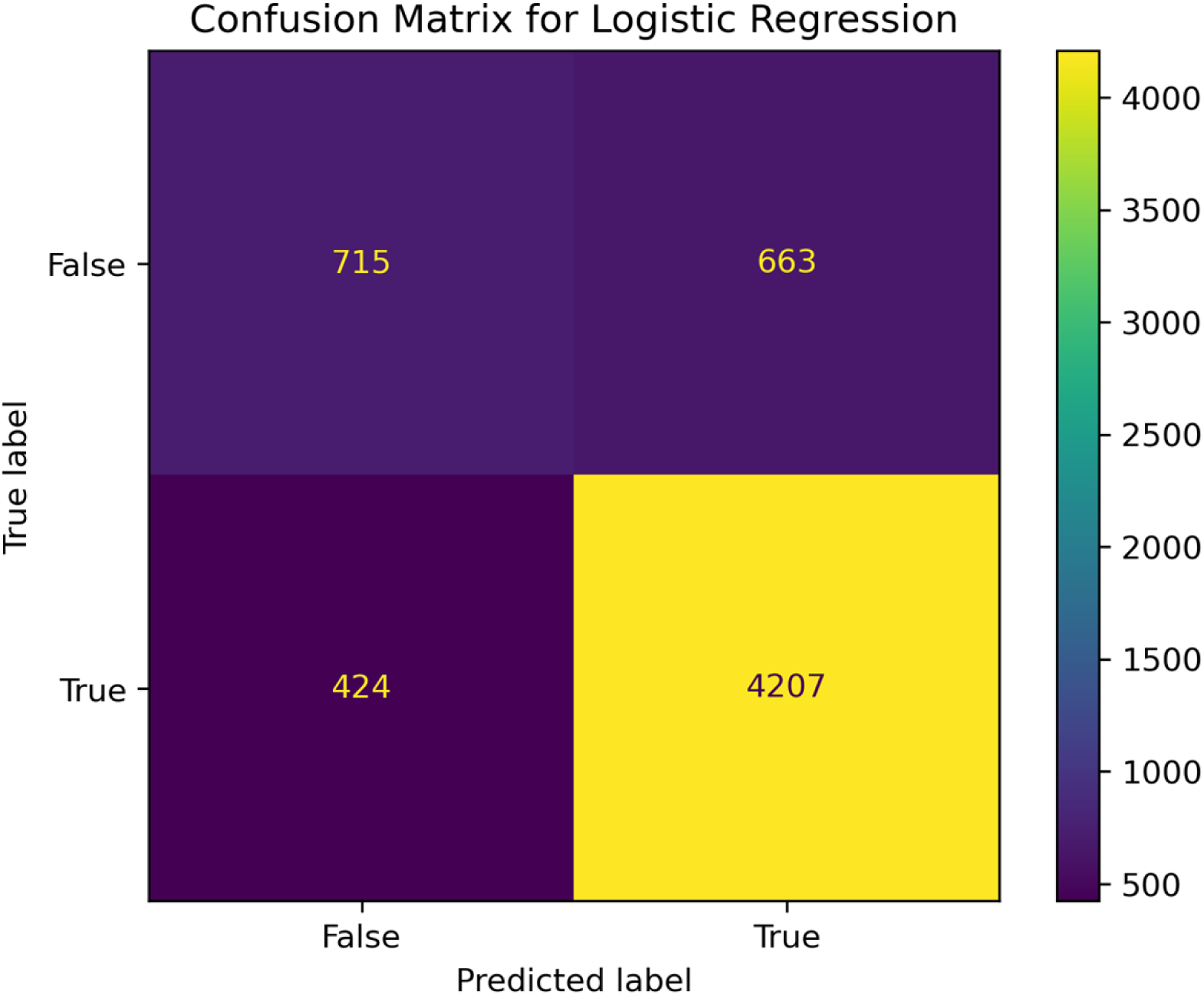
Confusion matrix for Logistic Regression model. True vs predicted classes for improved vs unimproved WASH access using Logistic Regression on test data.

### 3.5 Feature Importance and SHAP

The 4 most important features in XGBoost were: 1. Household wealth index, 2. Improved sanitation, 3. Place of residence, 4. Water at handwashing station.

SHAP showed higher wealth and urban residence pushed predictions toward “improved WASH”, while low wealth and rural residence pushed toward “unimproved WASH”.

**Fig 4.**
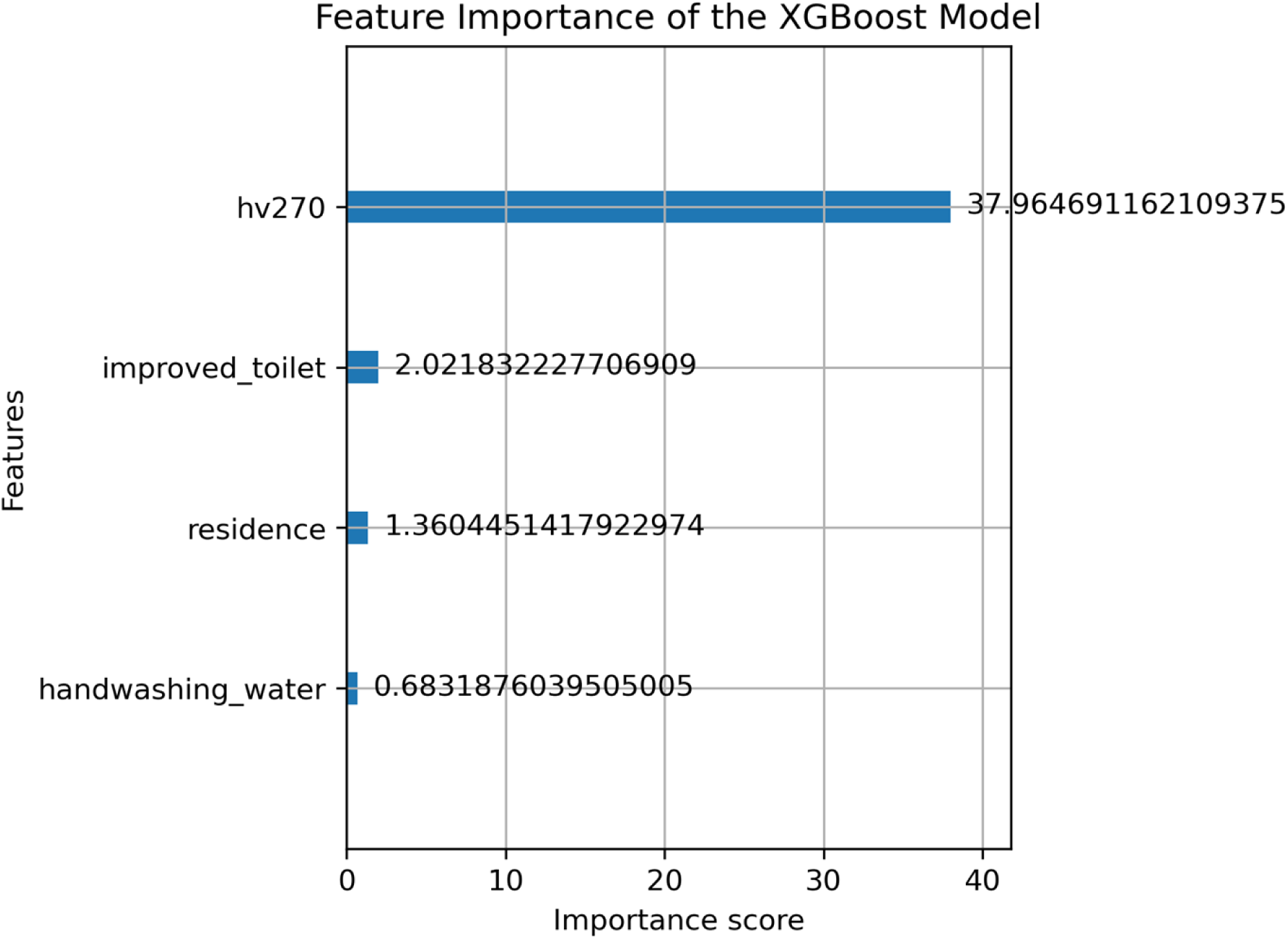
XGBoost feature importance ranking. Top predictors of improved household WASH access ranked by mean decrease in impurity. Household wealth index was the most important variable.

**Fig 5.**
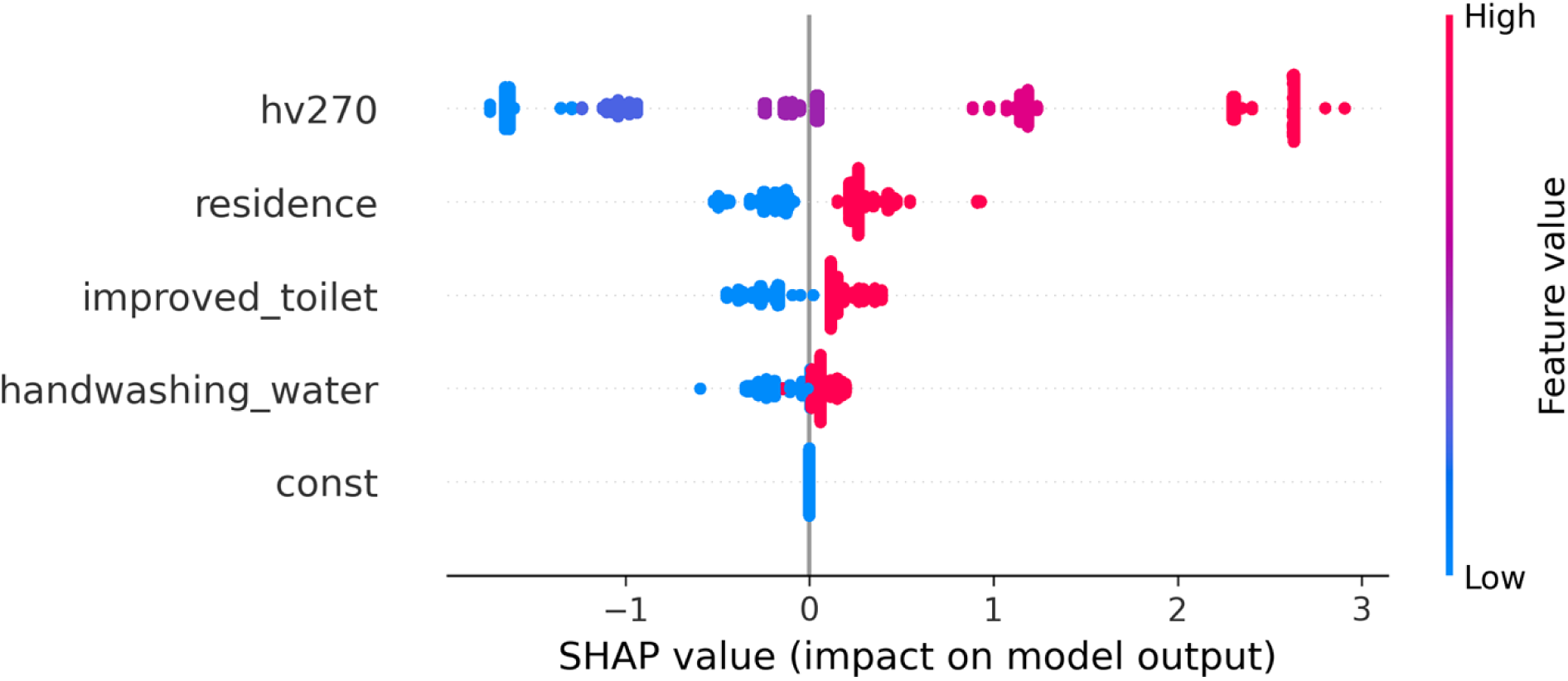
SHAP summary plot for XGBoost predictions. Each point represents one household. Position on x-axis shows impact on model output. Red = higher feature value, Blue = lower feature value.

**Fig 6.**
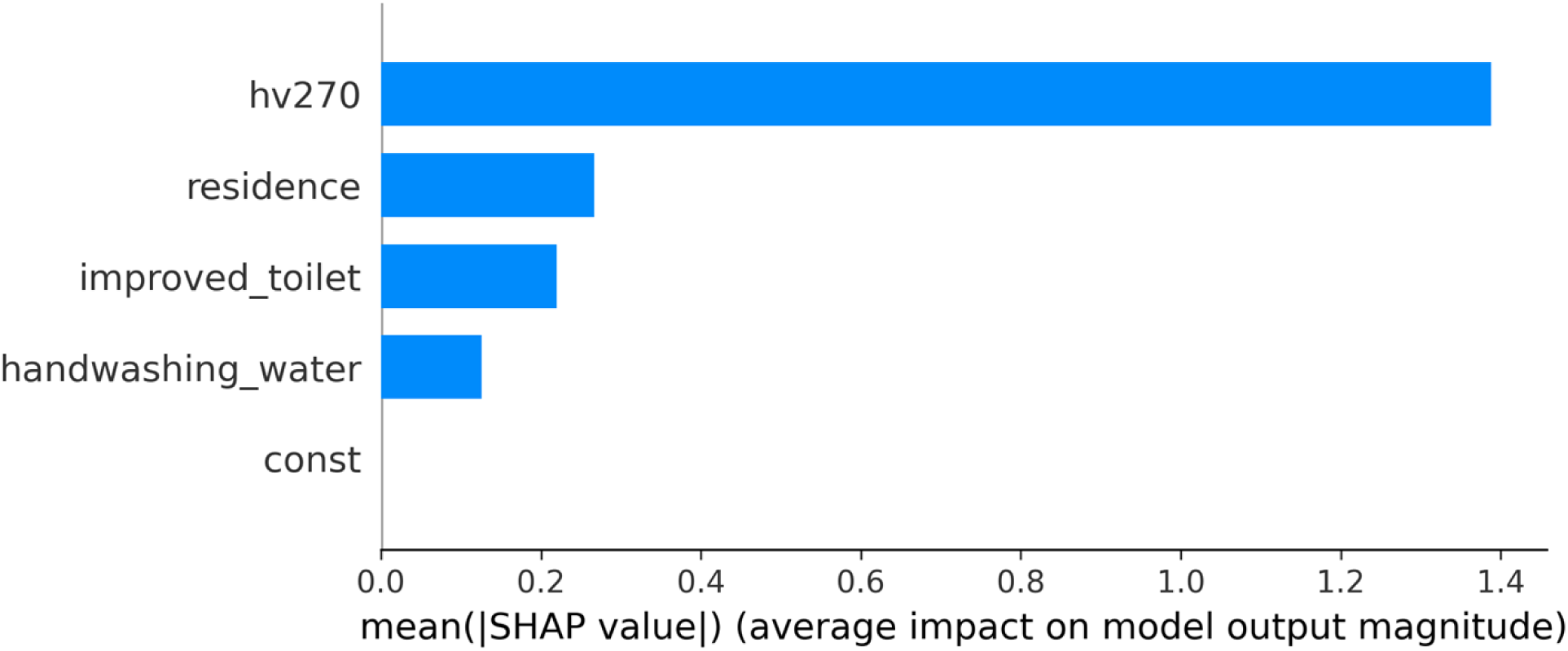
Mean absolute SHAP values for XGBoost predictors. Average impact of each feature on the model prediction. Higher values indicate greater importance to predicting WASH access.

### 3.6 Geospatial Risk of Unimproved WASH

To visualize geographic vulnerability, we aggregated predicted probability of unimproved household WASH to the 6 geopolitical zones. Figure 7 shows a clear South-North gradient. Southern zones had the highest average predicted probability of unimproved WASH: South West 0.91, South 0.88, and South East 0.87. Northern zones had lower risk: North Central 0.80, North West 0.74, and North East 0.72. Table 2 provides the zone-level averages.

**Table 4.** Average predicted probability of unimproved WASH by geopolitical zone, NDHS 2024.

| Zone | Average Risk |
| --- | --- |
| South West | 0.91 |
| South South | 0.88 |
| South East | 0.87 |
| North Central | 0.80 |
| North West | 0.74 |
| North East | 0.72 |

**Source: Author's computation from NDHS 2024 data.**

**Fig 7.**
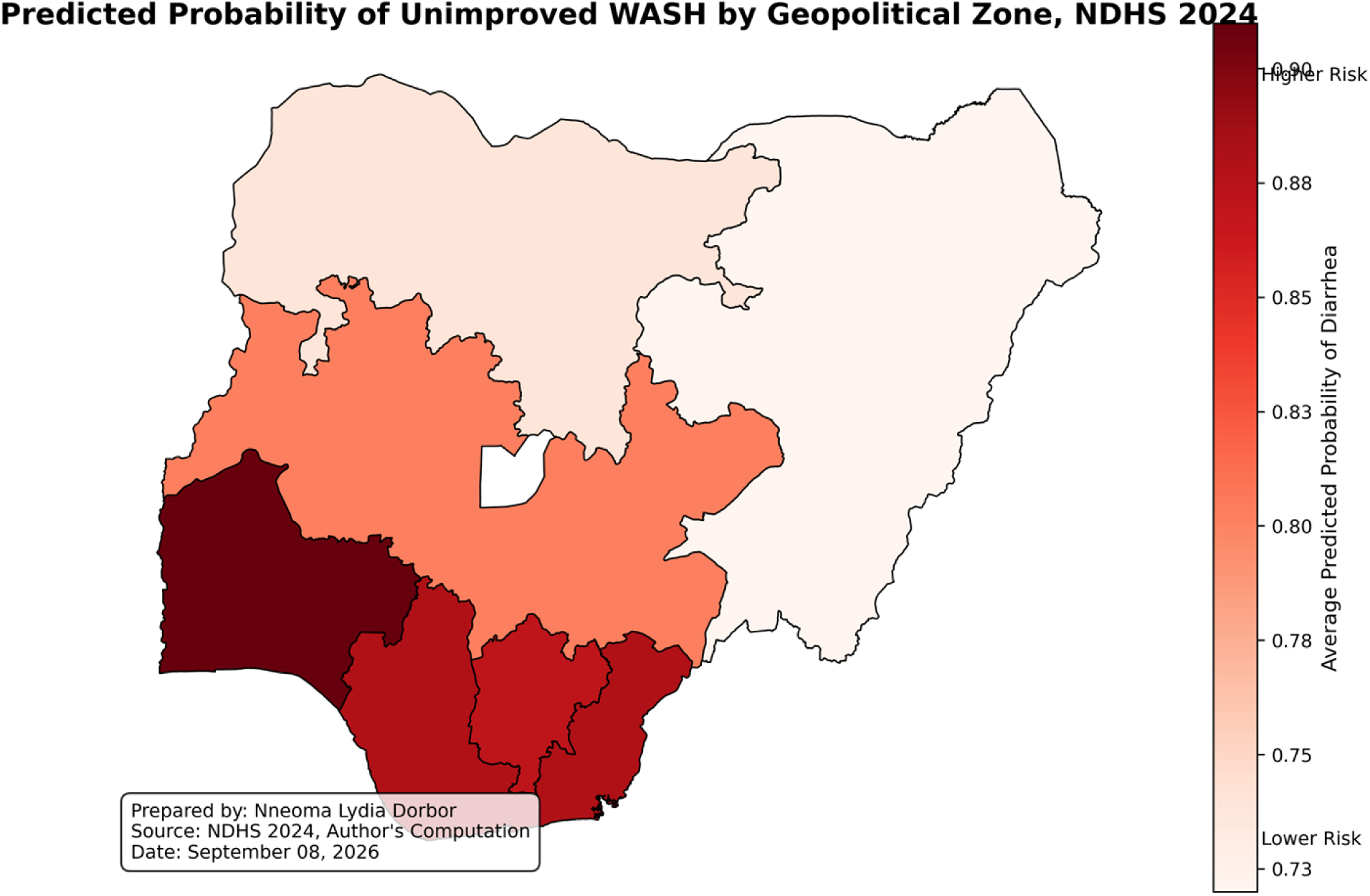
Predicted probability of unimproved WASH by geopolitical zone, NDHS 2024. Choropleth map showing average predicted probability of unimproved WASH aggregated to 6 geopolitical zones. Darker shades indicate higher risk. Southern zones had highest risk. Administrative boundaries sourced from HDX.

## 4. Discussion

This study compared four supervised ML algorithms for predicting household WASH access using recent nationally representative 2024 NDHS data. XGBoost achieved the strongest performance, though Random Forest was very close. This suggests that ensemble methods are well suited to capturing the non-linear relationships between wealth, geography, and WASH infrastructure.

The dominance of household wealth index as the top SHAP predictor reinforces that WASH inequity in Nigeria is fundamentally a socioeconomic problem.[1][3] Households with greater resources are more able to afford boreholes, improved latrines, and soap. The strong contribution of place of residence further highlights the infrastructure gap between urban and rural areas, particularly in northern Nigeria.

These findings have direct implications for NTD control. Many NTDs including soil-transmitted helminths and schistosomiasis are transmitted via poor WASH conditions.[7][17] By mapping vulnerability at the household level, ML can help NTD programs move from blanket distribution to targeted WASH + MDA interventions in high-risk communities.

While this study focused on WASH prediction, its relevance to AMR is indirect. We did not measure antimicrobial use or resistance. However, by reducing infection burden, improved WASH can reduce unnecessary antimicrobial demand, aligning with One Health strategies.[9][15] SHAP provided the critical transparency needed to make the “black-box” XGBoost model interpretable for policymakers.[10][11]

### 4.1 Policy Implications

1. **Target by wealth and geography**:

Prioritize integrated WASH funding for the poorest quintiles in rural North-West and North-East zones.

1. **Use predictive tools**:

State Ministries of Health could adapt this open-source model to identify LGAs most at risk for low WASH coverage.

1. **NTD-WASH Integration**:

NTD Master Plans should include WASH indicators from this model to guide resource allocation.

1. **Geographic targeting for diarrhea**:

The South West, South South, and South East zones showed the highest predicted probability of unimproved WASH. While overall WASH access is lower in the North, vulnerability appears concentrated in the South, likely due to flooding, urban density, and rainfall patterns. NTD and diarrheal disease programs should account for this South-North gradient.

### 4.2 Strengths and Limitations

**Strengths**:

Use of recent 2024 NDHS data; comparison of 4 ML models; use of multiple metrics beyond accuracy; full code reproducibility via GitHub; SHAP-based explainability.

**Limitations:**

1. Cross-sectional design prevents causal inference.
2. Limited to variables in NDHS - no water quality testing or geospatial data.
3. Internal 80:20 split only. External validation in future DHS rounds is needed.
4. Class imbalance: The outcome was imbalanced with 77% improved WASH. Future work could explore SMOTE or class weights to further improve recall for the minority class.
5. AMR and NTD outcomes were not directly measured.

## 5. Conclusion

Using 2024 NDHS data, we demonstrated that XGBoost with SHAP can accurately predict and explain household WASH inequities in Nigeria. Household wealth and rural residence were the key drivers. This interpretable ML framework provides actionable evidence to accelerate NTD elimination by targeting the most vulnerable populations. Future work should link these predictions to actual NTD prevalence and antimicrobial use data.

## Data Availability

The data underlying the results presented in this study are publicly available from the Demographic and Health Surveys (DHS) Program. Nigeria DHS data can be accessed at https://dhsprogram.com/data/ upon registration and approval of a data request. The minimal dataset used for analysis and the code used to generate the machine learning models and spatial maps are available in [Zenodo/GitHub repository name] at [DOI/link to be added upon acceptance].

https://github.com/nneoma-lydia/WASH-NDHS-Nigeria-2024

## Acknowledgments

We thank the National Population Commission of Nigeria, ICF, and the DHS Program for providing access to the 2024 NDHS. We also thank Prof. Gbenga Alege and Dr. Shitta Kefas Babale for their supervision and guidance.

## Author Contributions

Nneoma Lydia Dorbor: Conceptualization, methodology, data curation, formal analysis, visualization, writing—original draft.

Shitta Kefas Babale: Supervision, validation, writing—review & editing.

Gbenga Alege: Supervision, validation, writing—review & editing.

All authors approved the final manuscript.

## Declarations

### Ethics approval and consent to participate

This study used de-identified secondary data from the 2024 NDHS. Ethical approval for the original survey was obtained by the National Population Commission of Nigeria and ICF. Data access permission was granted by the DHS Program. No additional ethical approval was required for this secondary analysis.

## Consent for publication

Not applicable.

## Data availability

The 2024 NDHS dataset is available from the DHS Program upon approved request: https://dhsprogram.com/data. Analysis code is publicly available at: https://github.com/nneoma-lydia/WASH-NDHS-Nigeria-2024

## Supporting Information

### S1 Appendix. Analytical workflow for machine learning analysis of NDHS 2024 data

1. Data acquisition from 2024 NDHS household recode.
2. Data cleaning and preprocessing.
3. Variable selection.
4. Construction of binary improved-WASH outcome.
5. 80:20 training–testing split.
6. Model training: Logistic Regression, Decision Tree, Random Forest, XGBoost.
7. Evaluation: Accuracy, Precision, Recall, F1-score, AUC.
8. XGBoost feature importance and SHAP analysis.
9. Interpretation and visualization.

**S1 Table.** Abbreviations used in this study.

| Abbreviation | Meaning |
| --- | --- |
| AMR | Antimicrobial resistance |
| AUC | Area under the curve |
| DHS | Demographic and Health Survey |
| F1-score | Harmonic mean of precision and recall |
| JMP | Joint Monitoring Programme |
| ML | Machine learning |
| NDHS | Nigeria Demographic and Health Survey |
| NPC | National Population Commission |
| NTD | Neglected Tropical Disease |
| ROC | Receiver operating characteristic |
| SDG | Sustainable Development Goal |
| SHAP | SHapley Additive exPlanations |
| WASH | Water, sanitation and hygiene |
| WHO | World Health Organization |
| XAI | Explainable artificial intelligence |
| XGBoost | Extreme Gradient Boosting |

### S1 File

The trained XGBoost, Random Forest and Decision Tree models are available as Supporting Information file S1 at: https://github.com/nneoma-lydia/WASH-NDHS-Nigeria-2024.

